# An examination of the clarity of computerized cognitive training: Effect of instructions’ presentation mode on intrapsychic factors

**DOI:** 10.64898/2026.08.04.26359741

**Authors:** Christelle Nahas, Marc Gandit, Emmanuel Monfort

**Affiliations:** Translational Innovation in Medicine and Complexity (TIMC, UMR 5525), Université Grenoble Alpes, CNRS, Grenoble, France; Laboratoire InterUniversitaire de Psychologie (LIP/PC2S, EA4145), Université Grenoble Alpes, Grenoble, France

## Abstract

**Introduction:** Computerized cognitive training (CCT) is a promising and innovative solution to improve the quality of life for those experiencing age-related cognitive decline. The comprehension of instructions for CCT plays a crucial role in determining technology engagement. This study delves into the relationship between the presentation modes of CCT serious games instructions, their comprehension, and the resulting acceptability among older adults (aged over 65) without any known cognitive impairments.

**Methodology:** In a within-subjects experimental design, two types of CCT instructions were submitted to 128 older participants (mean age 71.5, 70% female): without visual cues and with visual cues. This approach was complemented by a study of the influence of self-efficacy and technology-related anxiety on the acceptability of the games.

**Results:** Instructions without salient visual cues were more acceptable for a complex functional game. Additionally, individuals with lower confidence in their cognitive abilities were less receptive to cognitive training, except for a highly familiar game.

**Conclusion:** The study highlights that older individuals may prefer simpler instructions for complex functional games, suggesting a preference for reduced cognitive load. It also shows the subtle role of self-efficacy in technology acceptance, except for the most familiar games, with higher cognitive self-confidence linked to greater acceptability. It emphasizes the importance of metacognition and self-efficacy in engagement when CCT involves mobilizing cognitive resources. It points the need for simple and personalized instructions to improve acceptance of CCT, and to contribute to the development of tailor-made interventions for older people.

## 1. Introduction

Amidst the growing concern over cognitive decline due to aging or neurodegenerative disorders, Computer Cognitive Training (CCT) is rapidly gaining popularity, particularly in rehabilitation contexts (1,2). This rising interest stems from the urgent need to confront the challenges associated with cognitive deterioration, a significant issue in contemporary healthcare (3,4). CCT’s ability to be tailored to individual needs, combined with its cost-effectiveness and ease of access, makes it an increasingly favored choice for enhancing cognitive functioning (5–7). As such, CCT represents a promising and innovative solution in the ongoing effort to improve the quality of life for those experiencing age-related cognitive decline. While some studies suggest that CCT can improve cognitive functions in older adults with cognitive impairments (6,8–10), the extent to which these improvements apply to everyday activities remains unclear (10–13). The challenge in assessing the transfer of cognitive gains to these activities may stem from various methodological issues. These include differences in the design of the studies (6,13–16). This contributes to the difficulty in determining how effectively cognitive improvements from CCT translate to practical, daily tasks.

A lack of engagement contributes to the lack of effectiveness of CCT, which potentially leads to poor rehabilitation outcomes (17–19). For older adults, the success of CCT is influenced by prior experience with computers, cognitive abilities, and the level of social support available (20). If the training is too intellectually challenging, there is an increased likelihood of participants dropping out. Therefore, simplifying computer use and providing additional assistance to those with cognitive impairments may lead to better adherence. Moreover, offering customized adjustments based on individual difficulties can promote effective engagement with CCT. Another vital aspect is the participants’ perception of the benefits and objectives of CCT. This encompasses their understanding and recognition of their cognitive deficits, which are key determinants in the utilization of CCT (20).

The concepts of user engagement and acceptance in technology are intricately connected (21–25). Acceptance involves the psychological aspects and the intention to use technology, regardless of previous experience (acceptability) or subsequent usage (acceptance) (25,26). Acceptability and engagement share influential factors such as perceived utility, self-efficacy, and outcome expectations, which are integral to acceptance models, also play a significant role in user engagement, as demonstrated in the rehabilitation model by Lequerica and Korte (17,27–29). Furthermore, usability, encompassing factors such as task completion time, user satisfaction, and ease of learning (25), is essential for fostering patient engagement, particularly in the development of cognitive training solutions for independent use. Poor usability can result in diminished engagement and adherence, adversely affecting perceptions of self-efficacy, perceived need, and outcome expectancy (30–33). Usability is closely linked to the quality of the instructions provided.

Key elements for successful engagement in rehabilitation, as pointed out by Lequerica and Kortte (17), include (1) the perception of rehabilitation needs, (2) the outcome expectancy for the treatment, and (3) the perception of self-efficacy, which play a big role in behavioral intention. These elements should be complemented by preparation such as goal setting and treatment planning. Additionally, the analysis of the experience and the reassessment of beliefs, attitudes, and expectations should be taken into consideration to maintain engagement in CCT. Clear and well-designed instructions facilitate the cognitive processing of information, thereby playing a significant role in enhancing comprehension (34–38). This aspect is especially vital for individuals with cognitive impairments, as research shows that systematically presented instructions can significantly improve learning and understanding of rehabilitation exercises (39,40).

Instructional design is pivotal in shaping information processing and, consequently, comprehension (34–41). Effective usability and acceptability (including perceived ease of use, self-efficacy, outcome expectancy) often rely on comprehension (17,25–27,30). Focusing on instructional design can help overcome cognitive and perceptual barriers associated with technology use by older adults, addressing issues like technology navigation, learning, information processing, memory, language (e.g., word choice), and visual needs (e.g., font size, colors) (41–45). Specifically for older adults, presenting information should involve simplicity, intuitive logic, moderate pacing, and minimal amount of irrelevant details. This approach is important because older adults often face challenges in processing information, memory, and visual perception, which can impede understanding instructions (36,42,45). Compared to younger individuals, older adults are more prone to having their attention captured by prominent visual cues, often struggling to inhibit reflexive shifts of attention towards these cues, even when instructed to ignore them or forewarned (46,47). This susceptibility can disrupt performance in cognitive training exercises when task-irrelevant exogenous cues are present (48). The design and attributes of these cues in cognitive training instructions critically influence how older adults respond, with factors like saliency, timing, and context playing a key role in attentional capture and task performance (49).

Beyond attentional capture, the mental effort required to process instructions is itself a determinant of comprehension and acceptability. According to Cognitive Load Theory (38), working memory has a limited capacity, and instructional materials that impose excessive intrinsic, extraneous, or germane load can hinder learning and task performance. For older adults, whose working memory capacity tends to decline with age, minimizing extraneous cognitive load (for instance by avoiding unnecessary or redundant visual elements) may be particularly important for successful engagement with CCT instructions. Salient visual cues added to reduce ambiguity could therefore have an ambivalent effect: while they may support comprehension when used sparingly, they could also increase extraneous load and overwhelm older users when combined with an already demanding task.

## 2. Material and methods

### 2.1. Study design

A within-subject design was used to assess the impact of salient visual cues on instructional comprehension and software acceptability in individuals over 65 without cognitive impairments, for CCT exercises, where each participant acts as their own control by experiencing all test conditions. This approach aimed to provide a comprehensive understanding of how instructional design and individual user characteristics interact to influence the effectiveness of cognitive training software for older adults. We presented two modes of CCT instruction using an online survey platform: Mode A without visual cues and Mode B with visual cues. We also examined the influence of participants’ cognitive self-efficacy and technology-related anxiety and self-efficacy on these factors. This design follows the method published by Nahas, Gandit, and Monfort (66), which used a within-subject design to evaluate the comprehension and acceptability of CCT serious games instructions among older adults.

Our hypotheses were twofold: we expected that instructions with salient visual cues (Mode B) would enhance comprehension and acceptability compared to instructions without such cues (Mode A), and that higher cognitive self-efficacy would correlate with greater acceptability, while higher technology anxiety and lower technology self-efficacy would be associated with lower levels of acceptability.

### 2.2. Participants

Adults aged 65 and older, fluent in French and without neurocognitive disorders, were recruited. They must have had access to a touchscreen tablet or computer but were discouraged from using phones for responding. A total of 128 participants were included in our study, with an average age of 71.5 years (SD = 5.16). Participant characteristics are presented in Table 1.

**Table 1.** Participants’ characteristics.

| Variables |  | N (%) |
| --- | --- | --- |
| **Gender** | Women | 90 (70.3 %) |
|  | Men | 38 (29.7 %) |
| **Marital Status** | Married | 60 (46.5 %) |
|  | Divorced | 28 (21.7 %) |
|  | In a relationship | 21 (16.3 %) |
|  | Widowed | 11 (8.5 %) |
|  | Single | 9 (7.0 %) |
| **Education Level** | PhD | 18 (14.0 %) |
|  | Masters | 35 (27.3 %) |
|  | Undergraduate level | 41 (31.8 %) |
|  | Baccalaureate degree | 25 (19.4 %) |
|  | Certificate of general education | 9 (7.0 %) |

### 2.3. Ethics statement

This study adheres to ethical guidelines for research involving human subjects, including compliance with European data protection regulations and the Declaration of Helsinki. It was approved by the university’s ethics board, the Comité d’Éthique pour les Recherches de Grenoble Alpes (CERGA), Université Grenoble Alpes (approval reference: CERGA-Avis- 2023-18). All participants read an information leaflet detailing the study’s objectives, procedures, and their right to withdraw at any time without consequence, and provided electronic informed consent by ticking a consent checkbox on the online survey platform prior to beginning the study.

### 2.4. Instructional design

The study selected six Cognitive Training (CCT) serious games, divided into two types: three analytical exercises and three functional exercises. The analytical exercises, which focus on specific cognitive skills like memory or attention, included “Barrage” (cancellation), “Le Bon groupe” (the right group), and “Memory.” The functional exercises, simulating everyday activities and targeting multiple cognitive capacities, were “La liste de course” (the shopping list), “Les courses” (shopping at the supermarket), and “Le GPS” (following a GPS route).

These exercises were grouped into two blocks: Block 1 for analytical exercises and Block 2 for functional exercises. Instructions for each exercise were divided into two main parts: the objective and the procedure. They were presented in two modalities: Modality A without visual cues, and Modality B with visual cues like arrows, colors, and shapes. Modality B was designed with reference to the rules of universal design which aim to create inclusive and user-friendly solutions (50)

The instructional materials, designed by the first author (CN), followed guidelines to facilitate information processing and reduce cognitive load. This included using multimedia, minimizing and breaking down information, highlighting relevant details, and using simple text and images (34–37,40,42,45,50,51). The main difference between the two modalities was the inclusion of salient visual cues in Modality B.

**Figure 1.**
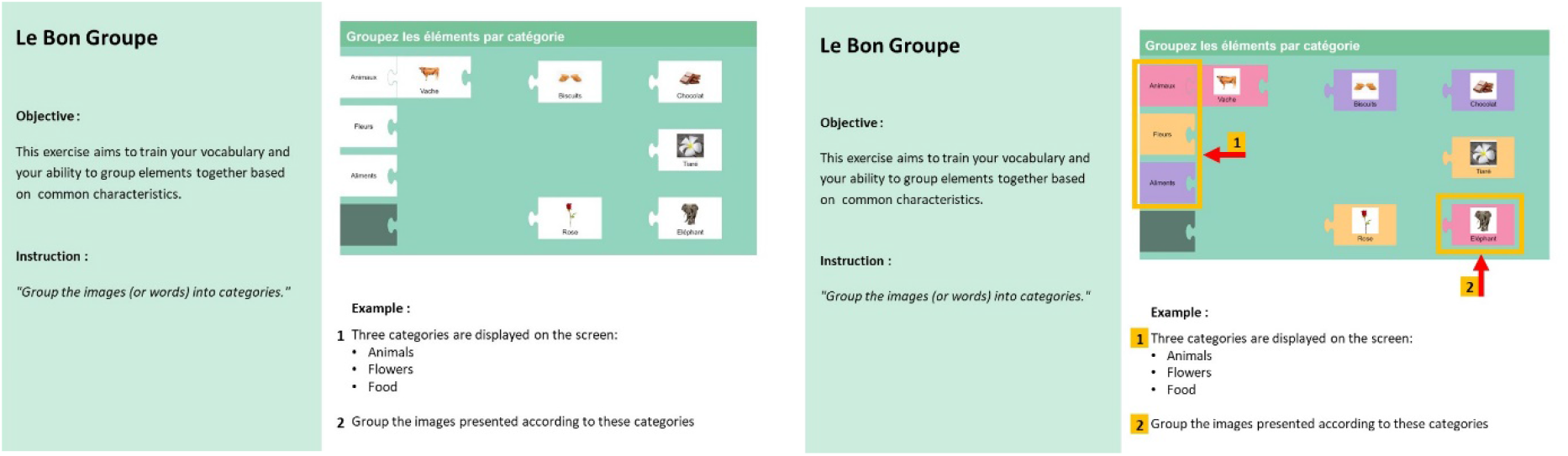
Example of instructions for “Le Bon Groupe”: Mode A without salient visual cues (left); Mode B with salient visual cues (right)

### 2.5. Measures

In this study, we assessed four main measures: acceptability, cognitive self-efficacy, technology self-efficacy, and technology anxiety.

Participants were asked to answer six questions on 5-point Lickert scales, after each of the visual instructions relating to the CCT serious games, in order to assess the participants’ degree of acceptance. These questions were based on the Reduced Instructional Materials Motivation Survey (52) to measure instructional comprehension. Initially in English, they were translated into French by a bilingual research team member. Furthermore, we incorporated questions about the perceived usefulness, expected outcomes, and self-efficacy. The questions were adapted from the French version of the eHealth acceptability scale (53), which is based on the technology acceptance model (27–29), already used with older people (54).

The Subjective Cognitive Complaints Questionnaire (SCCQ) was used to assess participants’ perception of their cognitive self-efficacy. It comprises 10 questions to which participants are asked to answer ‘yes’ or ‘no’ (55). Scores provide information on subjective cognitive functioning, but are not a cognitive screening tool.

An assessment of the overall sense of self-efficacy and anxiety in using technology is carried out using the self-efficacy scale developed by Compeau and Higgins (56), translated into French by Pelletier and Moreau (2009) (57). This scale includes 1) 10 items that ask the subject to determine if they think they can accomplish a task using a new computer application or new software, by answering ‘yes’ or ‘no’, and 2) 4 items that determine the anxiety related to using technology also answered by ‘yes’ or ‘no’.

### 2.6. Procedure

The study was conducted online. Data collection encompassed four main parts: a sociodemographic questionnaire, questions assessing self-efficacy and anxiety related to technology as well as the subjective perception of cognitive complaints, as well as the presentation of instructions for six CCT serious games in two modalities, in a counterbalanced way and an evaluation of instruction retention.

Each instruction for CCT serious games were followed by questions evaluating participants’ acceptability. Toward the end, participants’ retention of the instructions was evaluated through multiple-choice questions, ensuring a thorough assessment of their actual comprehension which is a guarantee of the quality of the measurements.

### 2.7. Statistical Analysis

Data were analyzed using the statistical package Jamovi 2.3.28. Given the non-parametric data distribution we chose to use non-parametric tests. A first series of analysis was performed to identify intergroup differences. For this purpose, we used Mann-Whitney U tests of median comparison. A second series of analysis was performed using *Spearman’s rank-order* correlations to measure the relationship between acceptability and understanding of the instructions of each of the exercises, cognitive self-efficacy, and self-efficacy and anxiety related to technology. Thirdly, we used linear regression models Multivariate to identify variables associated with acceptability score. The full models were compared with the parsimonious models using the likelihood ratio test to determine whether the inclusion of all the variables offered an additional advantage over the variables that had been found to be at least marginally significant in the bivariate analyses (*p* < .10).

## 3. Results

### 3.1. Differences in acceptability between visual modalities

In the conducted study, the acceptability of GPS exercise demonstrated a statistically significant higher average in Mode A (Mean = 19.8, SD = 2.75) compared to Mode B (Mean = 18.0, SD = 4.03), as indicated by the Mann-Whitney U test, U(1415), *p* < .01. No statistically significant difference was found in acceptability between modalities for the other exercises. Interestingly, despite initial hypotheses positing enhanced acceptability with salient visual cues in Mode B, the results generally favored Mode A, which lacked such cues. Detailed statistics of the paired-samples t-tests can be found in Table 2.

**Table 2.** Mann-Whitney U tests of median comparison between Modality A and B for the six games.

| Games | Mode A |  |  | Mode B |  |  | U | p |
| --- | --- | --- | --- | --- | --- | --- | --- | --- |
|  | n | M | SD | n | M | SD |  |  |
| Le Barrage | 64 | 19.6 | 2.29 | 65 | 20.0 | 2.39 | 1859 | .30 |
| Le Bon groupe | 68 | 20.1 | 2.42 | 61 | 19.0 | 2.72 | 1774 | .15 |
| Memory | 65 | 20.3 | 2.79 | 64 | 19.7 | 2.88 | 1874 | .33 |
| Le GPS | 66 | 19.8 | 2.75 | 63 | 18.0 | 4.03 | 1415 | < .01 |
| Les Courses | 60 | 18.9 | 2.74 | 69 | 19.3 | 3.28 | 1874 | .35 |
| Liste de courses | 69 | 19.8 | 3.16 | 60 | 19.3 | 3.29 | 1874 | .35 |

### 3.2. Relation between acceptability, technology self-efficacy, technology anxiety, and cognitive self-efficacy

Correlation analyses show a significant inverse correlation between the acceptability score and the score obtained on the SCCQ scale (ρ = -0.29, *p* < .001). A significant positive correlation was also observed between the anxiety score and the score obtained on the SCCQ scale (ρ = 0.24, p < .01). No statistically significant correlation was found between, on the one hand, the acceptability score and the score obtained for technological self-efficacy (ρ = 0.01, *NS*) and, on the other hand, the technological anxiety score (ρ = 0.02, *NS*).

### 3.3. Variance in acceptability explained by visual modalities and cognitive self-efficacy

Table 3 shows results of multiple linear regression for the acceptability scores for each exercise, highlighting the predictor variables’ contributions to acceptability. Preliminary analyses confirmed no violation of assumptions of normality. For the “Le barrage” exercise, the final model revealed a significant negative impact of the SCCQ score (*p* < .05), indicating that higher SCCQ scores (lower levels of cognitive self-efficacy) relates to lower acceptability. In contrast, technology self-efficacy and technology anxiety did not contribute significantly to the model. Similarly, in the “Memory” exercise, the SCCQ score exhibited a negative effect (*p* < .01), while technology anxiety showed a positive relationship (*p* < .05). Other variables did not demonstrate significant contributions.

**Table 3.** Parsimonious models with acceptability scores as outcomes.

| Game | Independent primary variables | Coefficient B | Std error | 95% confidence interval |  | t | p |
| --- | --- | --- | --- | --- | --- | --- | --- |
|  |  |  |  | lower | upper |  |  |
| Le barrage | Intercept <sup>a</sup> | 20.32 | 0.29 | 19.75 | 20.90 | 69.82 | < .001 |
|  | SCCQ | -0.26 | 0.11 | -0.47 | -0.04 | -2.33 | < .05 |
| Le bon groupe | Intercept <sup>a</sup> | 20.03 | 0.32 | 19.39 | 20.68 | 61.38 | < .001 |
|  | SCCQ | -0.12 | 0.12 | -0.36 | 0.12 | -0.97 | .33 |
| Memory | Intercept <sup>a</sup> | 20.50 | 0.35 | 19.80 | 21.19 | 58.53 | < .001 |
|  | SCCQ | -0.44 | 0.14 | -0.72 | -0.15 | -3.04 | < .01 |
|  | Anxiety for technology | 0.72 | 0.33 | 0.07 | 1.37 | 2.20 | < .05 |
| Le GPS | Intercept <sup>a</sup> | 20.39 | 0.50 | 19.41 | 21.37 | 46.76 | < .001 |
|  | Modality | -1.94 | 0.60 | -3.12 | -0.75 | -3.24 | < .01 |
|  | SCCQ | -0.38 | 0.16 | -0.70 | -0.05 | -2.32 | < .05 |
| Les courses | Intercept <sup>a</sup> | 19.96 | 0.37 | 19.21 | 20.70 | 53.69 | < .001 |
|  | SCCQ | -0.43 | 0.14 | -0.71 | -0.15 | -3.08 | < .01 |
| Liste de courses | Intercept <sup>a</sup> | 20.70 | 0.38 | 19.94 | 21.46 | 54.01 | < .001 |
|  | SCCQ | -0.60 | 0.15 | -0.89 | -0.32 | -4.16 | < .001 |

<sup>a</sup> Represents the reference level

In the case of the “GPS” exercise, acceptability was predicted by the visual modalities, mode B negatively impacting acceptability scores (*p* < .01). The SCCQ score also negatively predicted the acceptability score (p < .05). For “Les Courses,” the SCCQ score significantly negatively affected the acceptability score (*p* < .01). Other variables were non-contributory. In the “Liste de Courses” exercise, the SCCQ score also significantly and negatively influenced the acceptability score (*p* < .001), with no significant contributions from the remaining variables. Lastly, for “Le Bon Groupe,” the model indicated no statistically significant relationships between the predictors and the acceptability score.

## 4. Discussion

We aimed to explore the impact of salient visual cues within instructions on acceptability of CCT tools among older adults. Additionally, we aimed to investigate how the acceptability of instructional methods can be explained by the intrapsychic functioning of older people. The results obtained provide key insights contributing to the lack of acceptability of instructional information, offering a fresh perspective on the design and implementation of CCT. They also shed light on the significant relationship between cognitive self-efficacy and acceptability, contributing to better engagement in the use of CCT.

The main result of the study was that CCT instructions presented without salient visual cues were more acceptable than instructions with salient visual cues for one functional game (Le GPS), which contradicts conventional expectations. Such a preference aligns with the Cognitive Load Theory (38), suggesting that less complex interface reduced cognitive strain, thus enhancing acceptability. Conversely, salient visual cues, while intended to enhance clarity, may have increased cognitive load, overwhelming users with simultaneous information (42,44,45). This interpretation is consistent with recent applied work drawing on our earlier methodological paper (67), which underscores that the timing and modality of instruction presentation critically manage cognitive load and shape comprehension in digital learning environments for other populations, further supporting the idea that reducing extraneous load matters more than adding explanatory cues per se. This is further corroborated by a recent two-stage study on serious game interfaces in older adults with mild cognitive impairment (68), which found that game interfaces increased subjective cognitive load relative to non-game interfaces, and that ease of understanding and attentional focus—rather than the addition of explanatory features per se— were the pathways through which interface design reduced this load. However, the nature of the only game for which an effect was observed could have been perceived as more difficult than the other suggested games. Using a GPS is not a common part of older adult’s everyday life compared to going to the supermarket or creating a grocery list. This implies a learning process prior to using the game. It should be noted that this comparison was conducted separately across six games without correction for multiple comparisons; given this, the single significant modality effect observed for Le GPS should be interpreted with some caution and considered exploratory rather than confirmatory.

The second result, and the most consistent one across our models, is that individuals who feel less confident in their cognitive abilities might also be less receptive to accepting CCT games; unlike the modality effect described above, this association between cognitive self-efficacy (SCCQ) and acceptability emerged in five of the six regression models (Table 3), making it the most robust finding of this study. The perception of one’s cognitive capacities is a crucial aspect of the self-efficacy concept (21,22). When individuals perceive their cognitive abilities positively, they may exhibit greater confidence in handling intellectually demanding tasks. Older people thus show clear preferences for puzzle, strategy and educational games, which are the genres most frequently played (58). Confidence in games could enhance their overall self-efficacy, influencing willingness to take on challenges, persevere in the face of difficulties and make effective decisions. Thus, the perception of cognitive abilities appears to be central to the self-efficacy of older people, except for the game “Le bon groupe”, which required very little cognitive effort and adaptability. This also highlights the importance of metacognition, which is closely linked to self-efficacy, enabling individuals to understand and control their thoughts, strategies and performance (59). This would seem all the more important in the case of CCT, given that they are designed to train people at risk of cognitive decline (5–7). Metacognitive beliefs have a significant effect on the perception of cognitive tasks, particularly when seen as challenging (60,61). If individuals have negative beliefs about their cognitive ability, they are more likely to discontinue engagement in a task they assume to be difficult (62). Our results suggest that this relationship is confirmed for CCT: older individuals who acknowledge cognitive difficulties are less likely to readily accept cognitive training, despite being the ones presumed to need it. Implementing metacognitive interventions could therefore be an effective strategy to support cognitive training (63), except for the most familiar games.

It is worth noting that, contrary to expectations, technology anxiety was positively associated with acceptability of the “Memory” game. One possible explanation is that participants who felt anxious about technology may have appreciated this particular exercise precisely because its format and rules are widely familiar outside of a digital context, making the anxiety-provoking technological component less salient relative to the task itself; alternatively, this could reflect a degree of measurement noise given the exploratory nature of these game-specific models. This isolated finding invites replication before being interpreted further. More broadly, while our hypotheses anticipated a role for technology self-efficacy and technology anxiety alongside cognitive self-efficacy, neither variable was significantly correlated with acceptability at the bivariate level (see 3.2), and cognitive self-efficacy (SCCQ) emerged as the more consistent predictor across models. This suggests that, in this context, how older adults perceive their own cognitive abilities may matter more for CCT acceptability than their general relationship with technology.

While the study yields valuable insights, it’s important to acknowledge its limitations and exercise caution in interpreting the results. The use of a convenience sample restricts the generalizability of findings beyond the specific respondents involved. Future endeavors will involve extending this research to more specific population, for example, seniors engaged in preventive actions. Also, cognitive bias can impact acceptability scores (64). Consequently, further investigation through usability tests and qualitative interviews could offer additional insights into their perceptions and usage of CCT games.

In conclusion, our study showed that older people may prefer simpler, less visually complex instructions for complex functional games, and that cognitive self-efficacy significantly influences their acceptability, with the exception of the most familiar games. Further research involving usability tests and qualitative interviews could provide deeper insights into older adults’ perceptions and usage of CCT games. This suggests a complex interplay of factors influencing the potential adoption of these CCT games. These insights complement existing perspectives on designing CCT for older adults, emphasizing the need for personalized, user-centered approaches. This result might also be attributed to the absence of a co-design process, limiting the understanding of the target population’s needs (31). The results suggest that there is a need to explore other methods of helping older people get to grips with CCTs, such as the gradual, interactive introduction of visual cues or their presentation on demand of instructions for use by participants (using error-free learning techniques) (65). These approaches could potentially lead to improved acceptability rates.

## Author contributions

CN: Conceptualization, Methodology, Investigation, Formal analysis, Writing – original draft. MG: Conceptualization, Supervision. EM: Conceptualization, Methodology, Supervision, Writing – review & editing.

## Data Availability Statement

The data underlying the results presented in this study are not publicly available due to restrictions related to participant consent, which did not include authorization for open deposition of individual-level data. De-identified data are available from the corresponding author upon reasonable request.

